# Atypical functional connectivity of the left fusiform gyrus in infants at familial risk for developmental dyslexia

**DOI:** 10.1101/2022.02.24.22271455

**Authors:** Xi Yu, Silvina Ferradal, Jade Dunstan, Clarisa Carruthers, Joseph Sanfilippo, Jennifer Zuk, Lilla Zöllei, Borjan Gagoski, Yangming Ou, P. Ellen Grant, Nadine Gaab

## Abstract

**Importance:** Developmental dyslexia (dyslexia) is a genetic-based learning disorder affecting 7-10% of the general population and has detrimental impacts on mental health and vocational potential. Individuals with dyslexia show altered functional organization of the language and reading neural networks; however, it remains unknown how early these neural network alterations emerge in association with familial(genetic) vulnerability to dyslexia.

**Objective:** To determine whether the early development of large-scale neural functional connectivity is altered as a function of familial risk for dyslexia.

**Design:** This cohort study included 98 infants with (FHD+) and without (FHD-) a familial history of dyslexia recruited at Boston Children’s Hospital (BCH) between May 2011 and February 2019.

**Setting:** Participants underwent structural and resting-state functional magnetic resonance imaging in the Department of Pediatric Radiology at BCH.

**Participants:** FHD+ infants were defined as having at least one first-degree relative with dyslexia or reading difficulties and infants without familial risk for dyslexia (i.e., FHD-) were controls.

**Main outcomes and measures:** Whole-brain functional connectivity patterns associated with 20 pre-defined cerebral regions important for long-term language and reading development were computed for each infant. Multivariate pattern analyses were applied to identify specific functional connectivity patterns that differentiated between FHD+ and FHD-infants.

**Results:** The final sample consisted of 35 FHD+ (8.9 ± 2.4 months, 15 females) and 63 FHD- (8.3 ± 2.3 months, 36 females) infants. Multivariate pattern analyses identified distinctive functional connectivity patterns between the FHD+ and FHD-infants in the left fusiform gyrus (LFFG: accuracy = 0.55, *p*_*corrected*_ < 0.001, effect size: Cohen’s *d* = 0.76, 99% CI of the classification performance (classification accuracy-chance level) = [0.046, 0.062]). Moreover, the top five connections with greatest contribution to the classification performance connected LFFG with the frontal and temporoparietal regions of the language network.

**Conclusion and relevance:** The current study demonstrates that familial vulnerability to dyslexia is associated with an early onset of atypical functional connectivity of regions important for subsequent word form recognition during reading acquisition. Longitudinal studies linking the atypical functional network and school-age reading x(dis)abilities will be essential for further elucidating the ontogenetic mechanisms underlying the development of dyslexia.

**Key Points:** *Question:* Are functional topologies of language and reading-related regions in infancy associated with familial vulnerability to developmental dyslexia?

*Findings:* In a cohort study examining the resting-state functional connectivity of 98 infants during natural sleep, distinctive functional connectivity patterns of the left fusiform gyrus were observed between infants with and without a familial risk for dyslexia. These differences were evident despite comparable behavioral and socio-economic/environmental characteristics between the two groups.

*Meaning:* Familial vulnerability of dyslexia is associated with early alterations in the infant functional connectivity of key regions important for subsequent word recognition, suggesting an atypical neural scaffold for reading acquisition.

## Introduction

Learning to read words requires linking oral and written language [1], and therefore relies critically on the establishment of efficient communication among the language network comprised of left frontal and temporo-parietal regions and visual pathways located in the occipitotemporal cortex [2]. Despite exhibiting typical auditory and visual skills, approximately 7-10% of children fail to develop typical word reading and decoding abilities, termed developmental dyslexia (dyslexia, [3, 4]). The prevalence of dyslexia is higher among children with (FHD+) compared to those without (FHD-) a family history [5], suggesting a genetic basis of dyslexia.

Multiple dyslexia susceptibility genes have been identified [6, 7] and most of these have been hypothesized to impact early brain development (e.g., [8, 9]) potentially through atypical formation of cortical connectivity patterns underlying the foundations of reading acquisition [10, 11]. However, the specific neural mechanisms require further investigation [12]. Children with dyslexia are typically diagnosed after prolonged learning struggles (i.e., the “wait-to-fail” model [13]) and often exhibit persistent academic and psychosocial difficulties that negatively impact their mental health and vocational potential [14, 15]. Importantly, children at risk for dyslexia can be identified before reading onset [16] and interventions are more effective in young children due to heightened neuroplasticity [17, 18]. Therefore, it is critical to unravel the early developmental mechanisms explaining the etiology of dyslexia, which are essential for the development of strategies for early identification and intervention for children at risk.

Previous neuroimaging studies have revealed brain alterations associated with dyslexia. Individuals with dyslexia not only show neural deficits in regions important for language and reading [19-22], but also exhibit disrupted connectivity among these regions and their connections with domain-general cortices [23-27]. This suggests atypical neurobiological mechanisms in large-scale functional organization underlying the development of dyslexia. Importantly, both regional and connectivity deficits are evident before reading onset in children (i.e., pre-readers) who subsequently develop dyslexia [28-31], indicating that at least some of these observed neural alterations predate reading onset and are not a result of atypical reading experience.

Prospective longitudinal studies in FHD+ children with an increased dyslexia susceptibility provide a unique opportunity to examine the developmental trajectories of the neurobiology of dyslexia in relation to genetic risk. Atypical neural characteristics in language and visual processing areas are reported in FHD+ children before reading onset (e.g., [32-35]), as early as in infancy ([36, 37], see [13] for a review). While not all FHD+ children develop dyslexia [5, 38], they as a group often show lower reading skills than controls [39]. Moreover, neural alterations observed in FHD+ children are associated with subsequent reading skills [36, 37, 40, 41], suggesting the early onset of the atypical brain mechanisms underlying familial vulnerability to dyslexia. However, previous studies primarily focused on the early deficits in the local(regional) brain characteristics. Given the fundamental role of functional network mechanisms in typical and atypical reading acquisition, it is important to further examine whether the early development of large-scale functional connectivity might be altered as a function of familial risk for dyslexia.

Utilizing resting-state functional connectivity (FC) techniques, the current study examined the intrinsic functional connectivity in a group of 98 infants with and without familial risk for dyslexia. Resting-state FC measures the temporal synchronization of spontaneous brain activity using task-free functional MRI, permitting the characterization of the functional organization of the infant brain during natural sleep. We computed whole-brain FC patterns associated with each region important for long-term language and reading development. The same approach has identified prospective associations between infant FC and the language and foundational literacy development among children at school age [42], revealing a potential early neural scaffold for subsequent literacy development. Multivariate pattern analyses were applied to identify specific FC patterns that differentiated between FHD+ and FHD-infants. We hypothesized that regions playing important roles in subsequent language and literacy acquisition would show atypical FC patterns in infants with a familial risk for dyslexia compared to those without.

## Methods

### Participants

Ninety-eight infant participants (8.5 ± 2.3 months, age range: 4-13 months) were included in the current analyses (see recruitment details in SI method). Thirty-five infants (15 females) were classified as FHD+, defined as having at least one first-degree relative with dyslexia diagnosis (n=25) or reading difficulties (n=10), while 63 infants (36 females) were controls (i.e., FHD-). The two groups did not differ significantly in age or sex (all *p(ps)* > 0.1, Figure 2). No children had birth complications, neurological trauma, or developmental delays. The structural images of all infants were clinically interpreted as ‘normal’ by a pediatric neuroradiologist (P.E.G.). The current study was reviewed and approved by the institutional review boards at Boston Children’s Hospital and Harvard University. Before participation, informed written consent was obtained from a parent or legal guardian of each infant.

### Behavioral and environmental characterization

Infants’ developmental milestones in gross motor, fine motor, visual reception, and receptive and expressive language were successfully assessed in 29 FHD+ and 48 FHD-infants using the Mullen Scales of Early Learning assessment [43]. Raw scores were converted into standardized T-scores for subsequent analyses. Meanwhile, parents of all participants completed questionnaires documenting the home literacy environment (HLE, [44]) and socioeconomic status (SES), in terms of paternal education (Table S1). Potential group differences in the behavioral and the environmental measures were evaluated using two-sample *t*-tests and Wilcoxon–Mann–Whitney rank-sum tests, respectively.

### Imaging acquisition and preprocessing

We adopted the same acquisition parameters and preprocessing pipeline as applied in [42], and detailed information can be found in the supplementary method.

### Classification analyses on the functional connectivity patterns associated with long-term language and reading development

The preprocessed images of each infant were automatically parcellated into 78 cortical regions based on the 1-year-old Infant Brain Atlases [45] derived from the Automated Anatomical Labeling (AAL) atlas [46]. Given the study’s focus on the early neural mechanisms underlying reading development, seed regions were placed in the language and visual processing circuits, including inferior frontal (orbitofrontal, triangular, and opercular), precentral, temporoparietal (Heschl’s, inferior parietal, supramarginal, and angular gyri) and occipitotemporal (inferior temporal and fusiform) cortices (Figure 1A, e.g., [47-49]). Moreover, both hemispheres were considered given their joint involvement during reading activities in individuals at risk for [38] and diagnosed with dyslexia [19, 50], as well as early reading development [51]. The FC patterns of each seed were generated by correlating the BOLD timeseries between this region and each of the remaining cerebral cortices, which were Fisher transformed to Z scores for subsequent analyses (Figure 1B).

**Figure 1.**
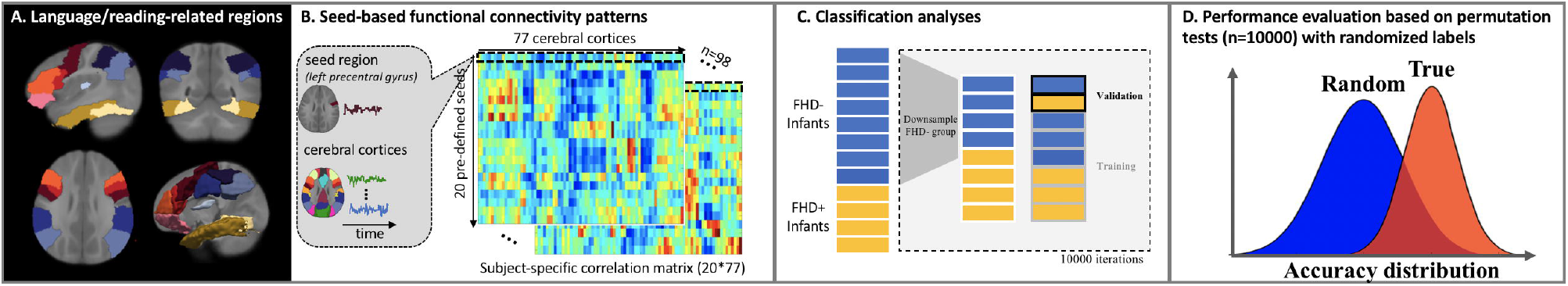
Flowchart showing the classification analyses for identifying distinctive functional connectivity (FC) patterns between FHD+ and FHD-infants. Panel A shows the slice views and 3D projections of 20 seed regions that were previously identified to be important for language and reading development. Panel B illustrates the computation process of the seed-based FC patterns for each infant. Specifically, region-specific time series were first estimated by averaging the BOLD signal across all the voxels in each of the cerebral cortices defined based on the 1-year-old Infant Brain Atlases (Shi et al., 2011) derived from the Automated Anatomical Labeling (AAL) atlas (Tzourio-Mazoyer et al., 2002). Seed-specific FC patterns were then generated by correlating the time series of each seed region (e.g., left precentral gyrus) with each of the other cerebral cortices, and the correlation coefficients were normalized to z-scores using Fisher’s transformation. Panel C presents the classification analyses applied to identify regions with distinctive FC patterns between FHD+ and FHD-infants. Note that a bootstrapping approach was adopted to generate a balanced sample of 35 FHD+ and 35 FHD-infants (randomly selected from the 63 FHD-subjects) for the classification analyses, which was repeated 10,000 times to reduce sampling bias. Panel D illustrates the evaluation of classification results against a null distribution derived from the permutation tests where the same classification analyses were performed but with randomized labels.

Classification analyses were performed using a Support Vector Machine (SVM) classifier based on the LIBSVM package [52]. The FC patterns of one seed were entered into the SVM analyses each time (Figure 1C). A leave-one-pair-out cross-validation strategy was used for tuning the SVM hyper-parameters and estimating the accuracy of the classifier. To deal with the imbalanced sample between the FHD+ and FHD-groups, a bootstrapping approach was further adopted where 35 samples were randomly selected from the FHD-pool to combine with the data of 35 FHD+ infants. This process was repeated 10,000 times to reduce sampling bias and generate a distribution of classification accuracies (i.e., the true distribution). To evaluate the classification performance, a permutation test was also conducted within each iteration, following the same procedure except that the FHD labels were randomly assigned, generating a null distribution over 10,000 iterations (Figure 1D). A paired *t*-test was performed to evaluate whether the true distribution was significantly higher than the null distribution. Moreover, a distribution of the classification performance was further computed for each seed by subtracting the classification accuracies of the permutation tests from those derived from the true labels within the same iteration. Upon this distribution, a 99% confidence interval (CI) was calculated to see if the lower CI bound was above 0, indicating better-than-chance-level classification performance (see a similar approach in [53]). Finally, for the seed-specific FC patterns with classification results significantly outperforming the permutation tests (i.e., *p*_*corrected*_ < 0.5) and demonstrating a positive CI, the effect size was further measured using Cohen’s *d*, where larger magnitudes indicate stronger classification performance. Only seed regions showing a medium or large effect (Cohen’s *d* > 0.5, [54]) were considered in the current study as exhibiting distinctive functional connectivity patterns between the FHD+ and FHD-groups. The classification sensitivity and specificity properties were also computed. Finally, for each region identified, we further estimated the weight information of every feature (i.e., FC between the seed and each cortical region), representing its contribution to the classification performance. To do this, path-specific weights were first calculated based on the bootstrapped samples (35 FHD+ and 35 FHD-infants) employed in each iteration, corrected for between-feature dependencies [55], and then averaged across the 10,000 iterations as the mean weight of each path.

## Results

### Psychometric and environmental characteristics

No significant differences were observed between the FHD+ and FHD-infants in any of the five developmental areas (*ps* > 0.05, Figure 2B). The two groups were also comparable in SES (parental education) and HLE characteristics (with the exception of higher frequency of rhyme and joke sharing among the FHD+ than the FHD-families, *p* = 0.02, uncorrected, see the full results in Table S1).

**Figure 2.**
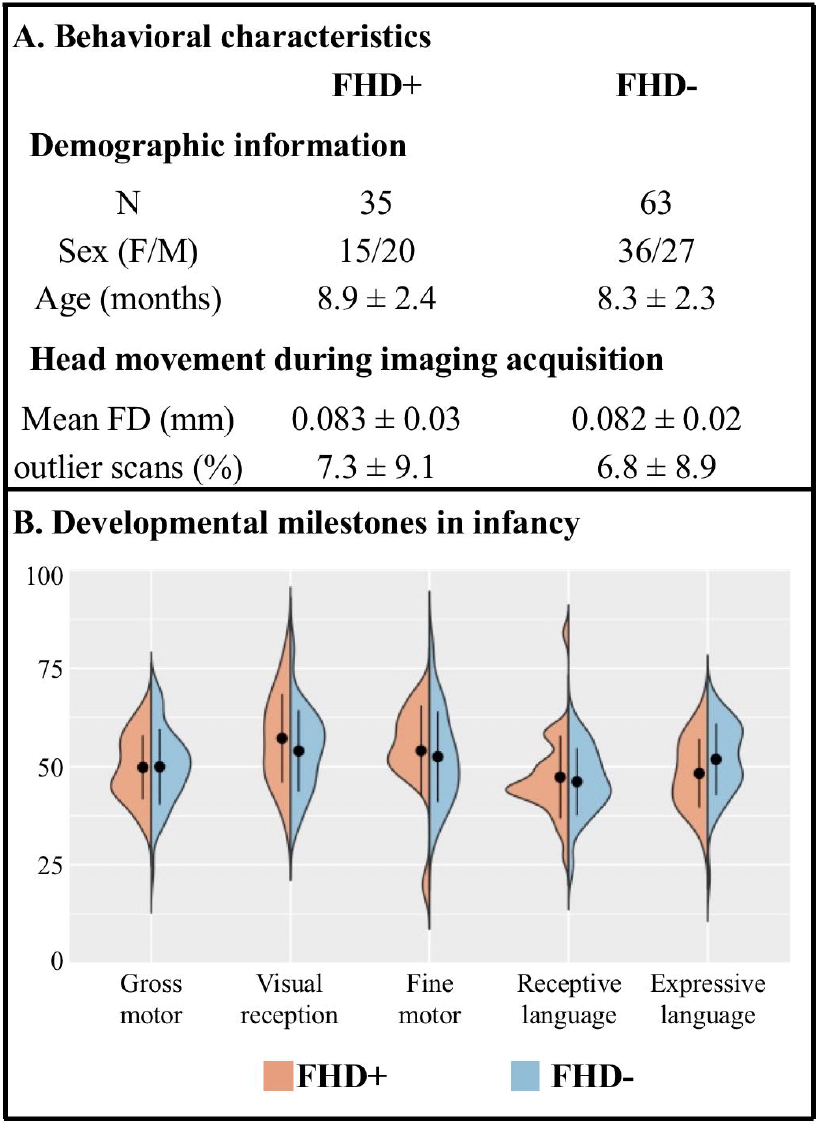
Demographic and behavioral characteristics of the FHD+ and FHD-infants. Panel A presents the demographic characteristics and head movement during scanning of the two groups. Panel B shows the behavioral development in all five areas for the two groups, as demonstrated by the T-scores (mean = 50, standard deviation = 10) of the Mullen Scales of Early Learning assessment. All direct comparisons yielded insignificant results (*ps* > 0.05).

### Classification results on the functional connectivity patterns

Head movement during imaging acquisition was similar between the FHD+ and FHD-infants, in terms of the percentage of outlier images and the mean framewise displacement after outlier removal (*ps* > 0.05, Figure 2A). Among the 20 seed regions associated with language and reading development, classification analyses identified distinctive functional connectivity patterns between the FHD+ and FHD-infants only in the left fusiform gyrus (LFFG: accuracy = 0.55, *p*_*corrected*_ < 0.001, 99% CI of the classification performance = [0.046, 0.062], Cohen’s *d* = 0.76, Figure 3, see Table S2 for the whole classification results). The classification performance based on the FC patterns of the LFFG also showed similar sensitivity (sensitivity = 0.54, *p*_*corrected*_ < 0.001, 99% CI of the classification performance = [0.037, 0.055], Cohen’s *d* = 0.56) and specificity (specificity = 0.56, *p*_*corrected*_ < 0.001, 99% CI of the classification performance = [0.054, 0.070], Cohen’s *d* = 0.76) characteristics. Finally, the weight of each path for this classification performance was computed and presented in the Figure 3C. The top five connections with greatest contribution to the distinctive FC patterns between the FHD+ and FHD-infants were paths connecting LFFG with the bilateral inferior parietal, right inferior occipital, left middle frontal, and right supramarginal gyri (see Table S3 for the full weight results).

**Figure 3.**
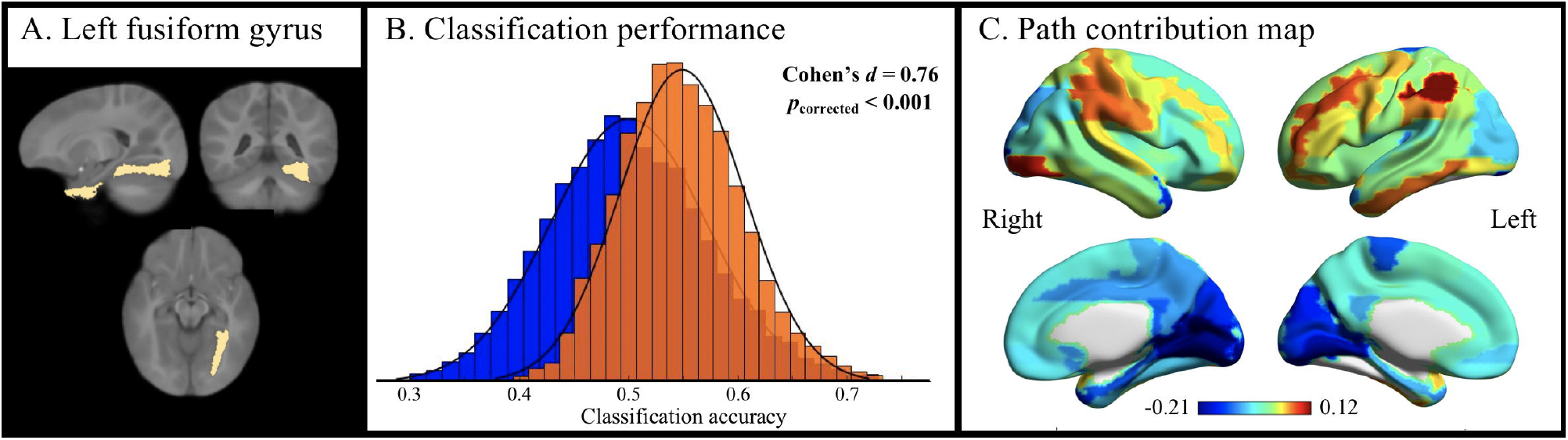
Distinctive functional connectivity patterns of the left fusiform gyrus (LFFG) between the FHD+ and FHD-infants. Panel A shows the slice views of the LFFG. Panel B illustrates the distribution of classification accuracies based on the true group labels (red) against the null distribution based on permutation results (blue). Panel C presents the contribution of each path to the familial risk classification performance by projecting the weight of each functional connectivity onto the corresponding cerebral cortex that LFFG connects to. Images were prepared using MRIcron and BrainNet software (Xia et al., 2011).

## Discussion

Here we demonstrate that the functional connectivity alterations associated with familial vulnerability of dyslexia are evident from as early as infancy. Specifically, early functional connectivity patterns of language- and reading-related regions were computed for a large cohort of 98 FHD+ and FHD-infants with well-balanced cognitive and critical environmental characteristics. Utilizing a machine learning approach, we identified distinctive functional connectivity patterns of the left fusiform gyrus (LFFG) between infants with and without familial risk for dyslexia. Moreover, the connections between the LFFG and the regions constituting the language network contributed the most to the FHD classification, with language network involving the frontal (left middle frontal gyrus) and temporoparietal regions (bilateral inferior parietal and right supramarginal gyri).

Distinctive FC patterns of LFFG between FHD+ and FHD-infants suggest that atypical network mechanisms associated with familial vulnerability to dyslexia emerge early in development. The fusiform gyrus is a key component of the ventral occipitotemporal cortex (VOTC) supporting high-level visual recognition of categories in adults, such as faces (fusiform face area, [56]) and scenes (parahippocampal place area, [57]). The visual word form area (VWFA), which exhibits specialization for visual word recognition has been repeatedly shown to emerge within the middle LFFG after reading onset in both children and adults, indicating experience-dependent functional specialization [58, 59]. Yet, FC between the VOTC (which houses the LFFG) and the left frontal and temporo-parietal components of the language network is already present in pre-readers [60] from as early as birth [61]. These early-emerging connections have been postulated to provide an initial neural scaffold for establishing associations between word forms and their phonological and semantic content over the course of learning to read [2, 62]. Besides its fine-tuning to orthographical properties, the LFFG is also sensitive to grapheme-to-phoneme mapping, which is essential for literacy acquisition [63, 64]. Moreover, the infant FC patterns of LFFG have recently been shown to be longitudinally associated with subsequent phonological skills in children at 6 years old after reading onset [42]. This highlights the long-term cognitive significance of the LFFG and its FC in the trajectory of literacy development. Within this framework, the current observation of atypical FC of the LFFG in infants with a familial risk for dyslexia might indicate early alterations in the neural network mechanisms underlying the integration of oral language and visual word processing. In line with this interpretation, the paths connecting the LFFG and oral language regions were found to contribute most to the FHD classification, which further supports proposed associations between familial vulnerability to dyslexia and atypical connectivity between the visual pathways and the language circuits evident in infancy. Finally, the atypical large-scale functional connectivity of the visual pathways in FHD+ infants also aligns with well-established FC disruptions of the VWFA in children with dyslexia [24, 65], and further suggests that atypical FC characteristics of dyslexia are unlikely to be *purely* driven by persistent reading difficulty. Overall, collective evidence suggests a possible developmental mechanism of dyslexia in the functional connectivity alterations associated with familial risk that are evident from infancy. Such alterations might disrupt the neural communication between the language and visual processing systems, upon which reading experiences build to enable successful reading acquisition.

The atypical FC patterns of the VOTC observed in FHD+ infants also shed light on the developmental mechanisms underlying dyslexia-associated neural alterations of the VWFA. Hypoactivation in response to written words has been repeatedly reported within the VWFA among individuals with dyslexia [22, 66]. Critically, reduced functional activation for letters were also identified in the VWFA vicinity among pre-readers who subsequently developed dyslexia [67], suggesting neural deficits contributing to the onset of dyslexia within the LFFG. Nevertheless, it still remains unclear what specific mechanism causes functional disruptions to letters in this area in pre-reading children. We have previously shown that the (structural) connectivity patterns of the VOTC in pre-readers at age 5 predated and predicted its functional tuning to words at age 8 after reading onset [68], supporting the notion that inter-regional connectivity may play a critical role in shaping the functional specialization process over the course of reading development [69, 70]. Following this hypothesis, it can be contemplated that the familial risk associated disruptions in the FC of the LFFG might serve as a developmental mechanism underlying characteristic VWFA alterations associated with dyslexia later in development. It is interesting to further note that, with no prior knowledge of the association specificity between the FC and functional specialization within the VOTC, it is possible that the atypical LFFG functional network in infancy might also disrupt the functional specialization for other visual categories. Indeed, atypical recognition performance and neural responses to faces were observed among individuals with dyslexia, although to a lesser extent [67, 71]. Future studies are warranted to empirically examine the hypothesized link between infants’ functional organization of the VOTC and subsequent functional specialization of the VWFA and the category-specific nature of this relationship to later literacy development.

Our study represents the first investigation to identify the early FC alterations associated with familial history of dyslexia in infancy long before a potential diagnosis can be given based on behavioral characteristics and school performance. Previous longitudinal studies utilizing event-related potentials have showed atypical neural responses to speech perception in FHD+ infants in association with their subsequent language and reading impairments [36, 37, 72], suggesting long-lasting effects of early brain and perceptual atypicalities. The current results extend these discoveries by further specifying disruptions in large-scale functional organization between regions important for long-term language/reading development that are associated with familial risk. We hypothesize that these FC alterations, despite being small in infancy, might be amplified during the postnatal brain expansion period within the first two years of life [73]. This, in turn, may impact long-term language and reading development, possibly contributing to the characteristic neural and behavioral alterations observed among older FHD+ children who develop dyslexia. Moreover, our group has also identified distinctive neural mechanisms in FHD+ pre-schoolers who later developed typical reading abilities despite neural alterations associated with familial risk, suggesting protective mechanisms facilitating positive outcomes among at-risk children [38]. While it remains unclear whether such protective mechanisms may be genetic-based or experience-driven [74], these findings open the possibility of developing neural resilience early in life among at-risk children. Thus, longitufinal follow-up is needed in future to uncover whether these early FC alterations interact with environmental factors and determine how these factors collectively shape the trajectory of reading development among at-risk children. Addressing these critical questions will not only help illuminate the neurodevelopmental trajectory of neural characteristics underlying dyslexia, but also has the potential to inform approaches to early diagnosis and intervention.

In conclusion, the current study provides the first evidence of atypical functional networks in infants with familial risk for dyslexia compared to those without despite comparable developmental milestones and socioeconomic/environmental factors. These alterations were observed in the infant functional connectivity of the left fusiform gyrus, an area that underlies long-term language and literacy development. Moreover, early disruptions were seen in connections linking LFFG to language regions and thus are hypothesized to play an important role in the functional specialization of the VWFA during reading acquisition. This body of work highlights the early onset of atypical functional topologies within the infant brain associated with a familial predisposition to dyslexia. Longitudinal tracking from infancy is critical for delineating how atypical functional networks in infancy may contribute to the prediction of subsequent reading (dis)abilities, and uncover genetic and environmental factors underlying this developmental trajectory. Advancing our understanding of the ontogenetic mechanisms underlying dyslexia sets a critical foundation for improving early identification and intervention practices.

## Data Availability

All data produced in the present study are available upon reasonable request to the authors

## Data availability

All data and codes used in these analyses can be shared with any research team upon request. Due to Institutional Review Board regulations at Boston Children’s Hospital at the time of consent, our data cannot presently be uploaded to a permanent publicly available archive. Nevertheless, data sharing can be achieved through a Data Usage Agreement. We are strongly committed to data sharing in order to facilitate outcome replication efforts.

## Acknowledgments

This study was supported by the Eunice Kennedy Shriver National Institute of Child Health and Human Development #R01HD065762-01 (awarded to N.G. and P.E.G), the National Natural Science Foundation of China #32100867 (awarded to X.Y.), the Fundamental Research Funds for the Central Universities (awarded to X.Y.), the Charles H. Hood Foundation (awarded to N.G.), and the Boston Children’s Hospital Pilot Grant (awarded to N.G.).

We sincerely thank all the families for their participation in this longitudinal study.

