## Supplementary materials for "Atypical functional connectivity of the left fusiform gyrus in infants at familial risk for developmental dyslexia"

Supplementary methods.

Participant recruitment.

All infants were selected from an ongoing longitudinal study at Boston Children's Hospital (BCH, now moved to Harvard University) that aims to characterize neural trajectories underlying language and reading development from infancy to school-age. A total of 118 participants have successfully completed resting-state functional and structural MRI sequences at infancy between 4 and 13 months old. Among them, 20 subjects were excluded from further analyses due to poor image quality (n=5), atypical brain anatomy (n=2), and excessive head motion during imaging sessions (n=13), resulting in the final set of 98 subjects (35 FHD+, 63 FHD-) with usable resting-state fMRI data included in the current analyses.

Imaging acquisition.

Infant participants were scanned during natural sleep (N. Raschle et al., 2012) on a Siemens 3T Trio scanner with a 32-channel adult head coil. High-quality structural images were acquired using a motion-compensated multi echo MPRAGE sequence with the following parameters: slice number = 176, TR = 2270 ms, TE1,2,3,4 = [1.66, 3.48, 5.3, 7.12] ms, flip angle = 7◦, TI = 1450 ms, field of view= 220 $mm^2$, voxel size =1.1×1.1×1.0 $mm^3$. An 8-minute blood-oxygen level dependent (BOLD) weighted imaging was further collected with two acquisition sequences: for data acquired earlier, the parameters were TR = 3000 ms, TE = 30 ms, flip angle = 60°, voxel size =3 x 3 x 3 $mm^3$; whereas an additional simultaneous multi-slice (SMS) imaging technique with a short TR (950 ms) was applied to the later collected images (the other parameters remained the same). The ratio of the FHD+ and FHD- infants were equivalent across two acquisition sequences ($X^2 = 0.11$, $p = 0.74$).

Resting-state fMRI preprocessing.

Preprocessing was conducted following an infant-specific preprocessing pipeline implemented in the FSL toolbox (Smith et al., 2004) (see details in (Yu et al., 2021)). Structural images were first skull-stripped and segmented into gray matter, white matter (WM), and cerebral spinal fluid (CSF). FMRI images were corrected for slice timing and head movement, and then normalized to the UNC 1-year old infant template (Shi et al., 2011) via the high-resolution structural image of the same infant using affine transformations. Based on the head movement parameters obtained during motion correction, framewise displacement (FD) was computed (Power, Barnes, Snyder, Schlaggar, & Petersen, 2012) using an in-house script (https://github.com/xiyu-bnu/infant_restingstate_prediction). Outlier volumes identified as FD > 0.3 mm were marked with one preceding and two subsequent frames to minimize the spreading effect due to temporal filtering or spin history. All included participants had at least 5-min usable volumes with an average of 7.0% of outlier images. Outlier images were then coded as a binary vector and submitted into linear regression with the six continuous motion regressors and mean CSF and WM signals estimated based on the subject-specific anatomical masks. After linear regression, images were temporally band-pass filtered (0.01-0.1Hz) and spatially smoothed (Gaussian filter, FWHM = 6mm). Finally, the fMRI time series with multi-slice acquisition were further temporally resampled to a TR = 3000ms in order to keep a consistent temporal resolution across the whole sample.

Table S1. Environmental characteristics for FHD- and FHD+ infants.

| Questions for environmental characterization | | Whole sample | | |
|---|---|---|---|---|
| | | FHD- (%) | FHD+ (%) | Group comparison (Mann–Whitney U test) |
| Home literacy environment | | | | |
| 1. Total number of parent/adult books in the home | 0-10 | 6.3 | 5.7 | $p = 0.53$ |
| | 11-50 | 15.9 | 11.4 | |
| | 51-100 | 19.0 | 22.9 | |
| | 101-200 | 20.6 | 22.9 | |
| | 201-300 | 17.5 | 5.7 | |
| | >300 | 17.5 | 28.6 | |
| | N/A | 3.2 | 2.9 | |
| 2. Total number of children's books in the home | 0-10 | 0.0 | 2.9 | $p = 0.44$ |
| | 11-50 | 30.2 | 25.7 | |
| | 51-100 | 22.2 | 34.3 | |
| | 101-200 | 22.2 | 14.3 | |
| | 201-300 | 7.9 | 11.4 | |
| | >300 | 14.3 | 8.6 | |
| | N/A | 3.2 | 2.9 | |
| 3. Age (in months) of child when first read to | Prenatal | 19.4 | 25.0 | $p = 0.59$ |
| | Less than one | 35.5 | 18.8 | |
| | 1-2 | 25.8 | 25.0 | |
| | 3-5 | 9.7 | 12.5 | |
| | 6-9 | 0.0 | 6.3 | |
| | 10 or more | 3.2 | 0.0 | |
| | N/A | 6.5 | 12.5 | |
| 4. Amount of time at home that someone reads to child (hours/week) | Less than one | 7.9 | 5.7 | $p = 0.39$ |
| | 1 | 19.0 | 22.9 | |
| | 2 | 19.0 | 14.3 | |
| | 3 | 19.0 | 28.6 | |
| | 4-5 | 19.0 | 5.7 | |
| | 6 or more | 7.9 | 14.3 | |
| | N/A | 7.9 | 8.6 | |
| 5. How often do family members teach the child to count? (times/week) | Never | 27.0 | 28.6 | $p = 0.48$ |
| | 1-2 | 14.3 | 25.7 | |
| | 3-4 | 14.3 | 20.0 | |
| | 5-6 | 4.8 | 0.0 | |
| | Daily | 23.8 | 20.0 | |
| | N/A | 15.9 | 5.7 | |

| | | | | |
|---|---|---|---|---|
| 6. How often do family members teach the child the alphabet? (times/week) | Never | 23.8 | 25.7 | $p = 0.28$ |
| | 1-2 | 20.6 | 37.1 | |
| | 3-4 | 14.3 | 8.6 | |
| | 5-6 | 6.3 | 0.0 | |
| | Daily | 19.0 | 17.1 | |
| | N/A | 15.9 | 11.4 | |
| 7. How often do family members read newspapers, books, or magazines? (times/week) | Never | 6.3 | 8.6 | $p = 0.082$ |
| | 1-2 | 9.5 | 11.4 | |
| | 3-4 | 14.3 | 2.9 | |
| | 5-6 | 6.3 | 22.9 | |
| | Daily | 58.7 | 51.4 | |
| | N/A | 4.8 | 2.9 | |
| 8. How often do family members write messages, notes, or lists? (times/week) | Never | 3.2 | 5.7 | $p = 0.11$ |
| | 1-2 | 3.2 | 5.7 | |
| | 3-4 | 4.8 | 20.0 | |
| | 5-6 | 12.7 | 5.7 | |
| | Daily | 73.0 | 60.0 | |
| | N/A | 3.2 | 2.9 | |
| 9. How often do family members write letters, cards, diaries, stories, or poems? (times/week) | Never | 9.5 | 14.3 | $p = 0.47$ |
| | 1-2 | 63.5 | 48.6 | |
| | 3-4 | 11.1 | 11.4 | |
| | 5-6 | 1.6 | 0.0 | |
| | Daily | 7.9 | 17.1 | |
| | N/A | 6.3 | 8.6 | |
| 10. How often do family members share rhymes or jokes orally with the child? (times/week) | Never | 7.9 | 2.9 | $p = 0.015$ |
| | 1-2 | 15.9 | 2.9 | |
| | 3-4 | 4.8 | 25.7 | |
| | 5-6 | 3.2 | 2.9 | |
| | Daily | 61.9 | 62.9 | |
| | N/A | 6.3 | 2.9 | |
| Socio-economic status | | | | |
| Maternal highest educational degree | 8th Grade or Less | 0.0 | 0.0 | $p = 0.35$ |
| | HS/GED | 4.8 | 11.4 | |
| | Associate Degree | 3.2 | 5.7 | |
| | Bachelor's Degree | 31.7 | 14.3 | |
| | Master's Degree | 41.3 | 42.9 | |
| | Doctorate or equivalent | 19.0 | 17.1 | |
| | N/A | 0.0 | 8.6 | |
| | 8th Grade or Less | 0.0 | 2.9 | $p = 0.32$ |

| | | | |
|---|---|---|---|
| Paternal highest educational degree | HS/GED | 11.1 | 17.1 |
| | Associate Degree | 4.8 | 8.6 |
| | Bachelor's Degree | 31.7 | 14.3 |
| | Master's Degree | 25.4 | 20.0 |
| | Doctorate or equivalent | 25.4 | 22.9 |
| | N/A | 1.6 | 14.3 |

For items with multiple choices, response frequency for each option (in percentage) was listed. Given the ordinal nature of parental response, group effects were examined using the Wilcoxon–Mann–Whitney two-sample rank-sum tests after excluding the "N/A" responses.

FHD-: infants without family history of dyslexia; FHD+: infants with family history of dyslexia

Table S2. Classification (CL) results for all 20 seed regions important for long-term language and reading development

| | Accuracy | | | Sensitivity | | | Specificity | | |
|---|---|---|---|---|---|---|---|---|---|
| | Mean | 99% CI (CL performance) | Cohen's *d* | Mean | 99% CI (CL performance) | Cohen's *d* | Mean | 99% CI (CL performance) | Cohen's *d* |
| Left Hemisphere | | | | | | | | | |
| Pars Orbitalis | 0.48 | [-0.021, -0.006] | | 0.47 | [-0.038, -0.021] | | 0.50 | [-0.007, 0.010] | |
| Pars Triangularis | 0.46 | [-0.045, -0.031] | | 0.48 | [-0.030, -0.014] | | 0.44 | [-0.062, -0.045] | |
| Pars Opercularis | 0.42 | [-0.085, -0.070] | | 0.44 | [-0.070, -0.054] | | 0.41 | [-0.010, -0.083] | |
| Precentral Gyrus | 0.46 | [-0.047, -0.033] | | 0.48 | [-0.031, -0.015] | | 0.45 | [-0.067, -0.050] | |
| Heschl's Gyrus | 0.50 | [-0.006, 0.007] | | 0.51 | [0.002, 0.019] | | 0.49 | [-0.017, 0.000] | |
| Inferior Parietal Gyrus | 0.46 | [-0.052, -0.038] | | 0.47 | [-0.043, -0.027] | | 0.44 | [-0.062, -0.046] | |
| Supramarginal Gyrus | 0.47 | [-0.032, -0.018] | | 0.48 | [-0.026, -0.009] | | 0.47 | [-0.040, -0.025] | |
| Angular Gyrus | 0.49 | [-0.017, -0.004] | | 0.49 | [-0.018, -0.0018] | | 0.48 | [-0.021, -0.003] | |
| Fusiform Gyrus | 0.55* | [0.046, 0.062] | 0.76 | 0.54 | [0.037, 0.055] | 0.56 | 0.56 | [0.054, 0.070] | 0.76 |
| Inferior Temporal Gyrus | 0.53* | [0.025, 0.040] | 0.49 | 0.51 | [0.002, 0.020] | 0.10 | 0.55 | [0.046, 0.064] | 0.76 |
| Right Hemisphere | | | | | | | | | |
| Pars Orbitalis | 0.49 | [-0.019, -0.005] | | 0.49 | [-0.020, -0.003] | | 0.49 | [-0.019, -0.001] | |
| Pars Triangularis | 0.48 | [-0.030, -0.015] | | 0.49 | [-0.019, -0.002] | | 0.47 | [-0.043, -0.026] | |
| Pars Opercularis | 0.50 | [-0.010, 0.006] | | 0.49 | [-0.011, 0.005] | | 0.50 | [-0.009, 0.009] | |
| Precentral Gyrus | 0.52* | [0.008, 0.023] | 0.24 | 0.49 | [-0.024, -0.007] | | 0.55 | [0.039, 0.056] | 0.61 |
| Heschl's Gyrus | 0.50 | [-0.007, 0.008] | | 0.47 | [-0.033, -0.016] | | 0.53 | [0.017, 0.034] | |
| Inferior Parietal Gyrus | 0.48 | [-0.038, -0.023] | | 0.48 | [-0.039, -0.022] | | 0.47 | [-0.038, -0.021] | |
| Supramarginal Gyrus | 0.51 | [-0.0031, 0.011] | | 0.53 | [0.016, 0.033] | | 0.49 | [-0.025, -0.0079] | |
| Angular Gyrus | 0.45 | [-0.063, -0.048] | | 0.46 | [-0.050, -0.033] | | 0.43 | [-0.078, -0.061] | |
| Fusiform Gyrus | 0.46 | [-0.042, -0.028] | | 0.48 | [-0.031, -0.013] | | 0.45 | [-0.055, -0.039] | |

| | | | | | | |
|---|---|---|---|---|---|---|
| Inferior Temporal Gyrus | 0.49 | [-0.016, -0.004] | 0.51 | [0.002, 0.020] | 0.47 | [-0.039, -0.022] |

*Note.* Among the 20 seed regions previously identified to be associated with language and reading development, only the left fusiform gyrus (highlighted in red) showed distinctive functional connectivity patterns between the FHD+ and FHD- infants, demonstrated by the above-chance-level classification accuracy, sensitivity and specificity with a median or large effect size (Cohen's d > 0.5), as well as positive 99% confidence intervals (CI) of the classification performance.

* Classification performance based on the real group information significantly (after correcting for multiple comparisons) outperformed the null results derived from the randomized labels ($p_{\text{corrected}} < 0.05$)

Table S3. Path weights in the familial risk classification of the functional connectivity patterns of the left fusiform gyrus

| Cerebral cortices | Weight (mean ± standard deviation) |
|---|---|
| Left inferior parietal lobule | 0.112 ± 0.050 |
| Right inferior occipital gyrus | 0.089 ± 0.041 |
| Left middle frontal gyrus | 0.069 ± 0.052 |
| Right inferior parietal lobule | 0.066 ± 0.046 |
| Right supramarginal gyrus | 0.063 ± 0.041 |
| Left inferior temporal gyrus | 0.060 ± 0.039 |
| Right postcentral gyrus | 0.055 ± 0.040 |
| Left supramarginal gyrus | 0.052 ± 0.044 |
| Left orbitofrontal cortex (middle) | 0.049 ± 0.048 |
| Left precentral gyrus | 0.042 ± 0.042 |
| Left temporal pole (superior) | 0.039 ± 0.054 |
| Right middle frontal gyrus | 0.034 ± 0.051 |
| Right temporal pole (superior) | 0.033 ± 0.052 |
| Right superior temporal gyrus | 0.031 ± 0.051 |
| Left inferior occipital gyrus | 0.029 ± 0.041 |
| Left inferior frontal gyrus (triangular) | 0.028 ± 0.053 |
| Right orbitofrontal cortex (middle) | 0.024 ± 0.050 |
| Right precentral gyrus | 0.020 ± 0.045 |
| Right inferior temporal gyrus | 0.010 ± 0.040 |
| Right superior parietal gyrus | 0.010 ± 0.051 |
| Left angular gyrus | 0.009 ± 0.045 |
| Left superior parietal gyrus | 0.004 ± 0.062 |
| Left inferior frontal gyrus (opercular) | 0.004 ± 0.047 |
| Left superior frontal gyrus (dorsal) | 0.001 ± 0.047 |
| Left postcentral gyrus | 0.000 ± 0.043 |
| Left orbitofrontal cortex (inferior) | -0.001 ± 0.049 |
| Left middle temporal gyrus | -0.003 ± 0.051 |
| Left orbitofrontal cortex (superior) | -0.004 ± 0.043 |
| Right Heschl gyrus | -0.006 ± 0.0481 |
| Left insula | -0.007 ± 0.049 |
| Right rolandic operculum | -0.008 ± 0.042 |

| | |
|---|---|
| Right orbitofrontal cortex (superior) | -0.011 ± 0.048 |
| Right middle temporal gyrus | -0.011 ± 0.045 |
| Right inferior frontal gyrus (triangular) | -0.028 ± 0.050 |
| Left supplementary motor area | -0.028 ± 0.042 |
| Right superior frontal gyrus (dorsal) | -0.032 ± 0.044 |
| Left olfactory | -0.033 ± 0.051 |
| Right orbitofrontal cortex (inferior) | -0.033 ± 0.048 |
| Right insula | -0.034 ± 0.048 |
| Left middle cingulate gyrus | -0.036 ± 0.037 |
| Left rolandic operculum | -0.036 ± 0.043 |
| Right middle occipital gyrus | -0.037 ± 0.047 |
| Left superior frontal gyrus (medial) | -0.038 ± 0.040 |
| Left superior temporal gyrus | -0.038 ± 0.049 |
| Left anterior cingulate gyrus | -0.040 ± 0.043 |
| Left precuneus | -0.041 ± 0.057 |
| Right supplementary motor area | -0.044 ± 0.045 |
| Right anterior cingulate gyrus | -0.045 ± 0.045 |
| Right inferior frontal gyrus (opercular) | -0.045 ± 0.043 |
| Right superior frontal gyrus (medial) | -0.050 ± 0.042 |
| Left rectus gyrus | -0.051 ± 0.041 |
| Right rectus gyrus | -0.054 ± 0.044 |
| Right angular gyrus | -0.055 ± 0.041 |
| Right orbitofrontal cortex (medial) | -0.060 ± 0.040 |
| Left superior occipital gyrus | -0.062 ± 0.062 |
| Right fusiform gyrus | -0.069 ± 0.038 |
| Left orbitofrontal cortex (medial) | -0.070 ± 0.038 |
| Left middle occipital gyrus | -0.070 ± 0.052 |
| Left posterior cingulate gyrus | -0.081 ± 0.0423 |
| Right paracentral lobule | -0.081 ± 0.049 |
| Left paraHippocampal gyrus | -0.081 ± 0.046 |
| Right superior occipital gyrus | -0.087 ± 0.063 |

| | |
|---|---|
| Right precuneus | -0.089 ± 0.057 |
| Right middle cingulate gyrus | -0.092 ± 0.042 |
| Right olfactory | -0.093 ± 0.050 |
| Left temporal pole (middle) | -0.096 ± 0.048 |
| Left Heschl gyrus | -0.099 ± 0.047 |
| Right posterior cingulate gyrus | -0.121 ± 0.042 |
| Right paraHippocampal gyrus | -0.122 ± 0.046 |
| Left paracentral lobule | -0.124 ± 0.053 |
| Left calcarine cortex | -0.143 ± 0.060 |
| Right cuneus | -0.145 ± 0.0622 |
| Right temporal pole (middle) | -0.145 ± 0.049 |
| Left cuneus | -0.158 ± 0.059 |
| Left lingual gyrus | -0.192 ± 0.043 |
| Right calcarine cortex | -0.207 ± 0.052 |
| Right lingual gyrus | -0.210 ± 0.045 |
